# Full BLOOD count TRends for colorectal cAnCer deteCtion (BLOODTRACC): external validation of colorectal cancer prediction models in English primary care

**DOI:** 10.1101/2025.11.26.25341055

**Authors:** Pradeep S. Virdee, Jacqueline Birks, Tim Holt, Kym I.E. Snell, Gary Abel, Brian D. Nicholson

## Abstract

**Introduction:** Colorectal cancer has low survival rates when diagnosed late-stage. We previously developed sex-specific dynamic risk prediction models utilising trends in the full blood count (FBC), a blood test commonly performed in primary care, to support early detection. We aimed to externally validate these prediction models.

**Methods:** We performed a hybrid case-control and cohort study of patients with at least one FBC test. We first excluded FBCs within two years before diagnosis (cases) or study exit (controls) and selected the most recent FBC as the baseline test per patient from the resulting data. Patients were aged at least 40 years at baseline and had no history of colorectal cancer. The models included age (years) at baseline and simultaneous trends over historical haemoglobin, mean corpuscular volume (MCV), and platelet measurements measured over five years before baseline to inform two-year risk of colorectal cancer diagnosis. Performance measures included the c-statistic and calibration slope.

**Results:** We included 2,956,977 males and 3,561,349 females, with 0.4% (n=12,578) and 0.3% (n=11,939) diagnosed with colorectal cancer, respectively. The c-statistic (95% CI) was 0.73 (0.72-0.73) for males and 0.74 (0.74-0.75) for females. The calibration slope (95% CI) was 0.92 (0.89-0.94) for males and 0.95 (0.93-0.98) for females. Calibration was good in subgroups of patient data, except under-predicted risk in those aged 70+ years, White individuals, and those with higher IMD. The c-statistic (95% CI) was similar regardless of the number of FBCs used to define trend and increased as the longitudinal trend window increased until around 2.5-3.0 years for men (0.73 (0.71-0.74)) and 3.0-3.5 years for women (0.73 (0.72-0.75)) and decreased with increasing longitudinal windows thereafter.

**Conclusion:** Utilising temporal changes in the FBC test could enhance risk stratification for colorectal cancer. Further research may highlight approaches for improving predictive performance further.

## Introduction

Colorectal cancer is the fourth most common type of cancer[1] and second most common cause of cancer-related death[2] in the UK. Survival is associated with tumour stage at diagnosis: five-year survival is 90% at Stage I and 10% at Stage IV[3]. Around 55% of staged cases are diagnosed late-stage[4]. Earlier detection would improve the likelihood of successful treatment and reduce mortality[5]. Most diagnoses (55%) are made following urgent general practitioner (GP) referral for symptoms, as recommended by the National Institute for Health and Care Excellence (NICE)[6]. Many symptoms are common and non-specific, so many more referrals are made than cancers diagnosed, placing significant pressure on healthcare resources, particularly on colonoscopy capacity.

The full blood count (FBC) blood test measures 20 components of the blood and is commonly performed in primary care[7]. Abnormalities in many individual components, such as raised platelets/thrombocytosis, are associated with increased risk of colorectal cancer[8]. We previously reported statistically significant association between trends over repeat FBCs for most components and colorectal cancer diagnosis, with cases having on average a different trend within four years prior to diagnosis compared to cancer-free patients[9]. We found that relevant trends may appear before abnormal FBC thresholds[10–12] that prompt further cancer investigation[13–15] are reached, showing potential for earlier diagnosis.

We developed the BLOODTRACC models, sex-stratified diagnostic prediction models derived using joint modelling and English Clinical Practice Research Datalink (CPRD) GOLD primary care data[16]. Each model utilises age and trends in haemoglobin, mean corpuscular volume (MCV), and platelets to determine two-year risk of colorectal cancer. Internal validation found a c-statistic of 0.76 for females and 0.75 for males, using data measured earlier than two years before diagnosis. We aimed to externally validate the BLOODTRACC models to assess performance in external primary care data.

## Methods

Study reporting follows the TRIPOD guidelines[17]. Ethical approval was provided by the CPRD Research Data Governance on behalf of the Health Research Authority (protocol number: 22_001798). Data preparation and analysis was performed in RStudio (R V4.1.3).

### Study population

Patient data was from the English CPRD AURUM primary care electronic health record database, available for patients registered between 1^st^ January 2000 and 31^st^ December 2018. The data was linked to the National Cancer Registration and Analysis Service (NCRAS) Cancer Registration, Office for National Statistics (ONS) Death Registration, Hospital Episode Statistics (HES) Admitted Patient Care, Outpatient, and Accident and Emergency, and Small Area Level Data. SNOMED-CT, Medcodes, and ICD10 codes for each data item are available at https://github.com/PradeepVirdee/BLOODTRACC_ModelValidation_CPRDAurum[18].

Patients were aged at least 40 years with at least one haemoglobin, MCV, and platelet measurement available. Patients registered with their primary care practice for less than one year, with a history of colorectal cancer before baseline (defined below), or ineligible for linkage to NCRAS, HES, and ONS were excluded.

### Study design

We used the same design as the model derivation study. We performed a hybrid case-control and cohort study. First, we identified the date of diagnosis in cancer cases and study exit date in cancer-free controls and excluded haemoglobin, MCV, and platelet measurements within the two years prior, ensuring we used data measured at a sufficiently earlier phase that increases the likelihood of successful clinical intervention to improve prognosis. From the resulting dataset, the baseline test was the most recent test available. Tests performed before baseline were considered historic. Trends were identified using all historical tests available up to five years prior to the baseline test. Risk predictions are therefore made from the baseline time-point, incorporating information from historical tests. A five-year longitudinal period was chosen based on our previous work showing differences in trends between patients with and without a diagnosis confined to five years pre-diagnosis[9]. A graphical depiction of this study design has previously been reported[16].

### Outcome

Due to the two-year exclusion window described above, there were no diagnoses or censored patients within the two years following the baseline test. Therefore, the outcome was a diagnosis of colorectal cancer at two years (+/-three months to allow for a time-to-event distribution to form) after the baseline test. Patients without a diagnosis at two years (+/-three months) post-baseline were censored at the earliest of date of leaving the practice, death, 31^st^ December 2018, or two years after their baseline test. Diagnoses were identified from the NCRAS database using ICD10 codes C18-C20.

### Covariates

The models rely on age (years) at baseline test, sex, and trends in historic haemoglobin, MCV, and platelet measurements up to baseline test. Date of birth and sex were provided for all patients by the CPRD. We cleaned blood test results to remove values outside biologically plausible ranges, which have previously been reported[9], and standardised to the same unit of measurement following guidance from previous work[19]. There was little (<~5%) missing haemoglobin, MCV, and platelet data among all FBCs so no data imputation was performed. Instead, the derived models employed joint modelling, which includes a mixed-effects modelling component that can account for sporadically measured and unbalanced data, so every blood test result, as available, was included in this validation study.

### Model performance

Overall performance was assessed using the Brier score[20]. Discrimination was assessed using the c-statistic and Royston and Sauerbrei’s D-statistic[21]. Calibration was assessed using the calibration slope and calibration plots. Calibration plots were derived by first categorising patients into 20 equally sized groups of predicted two-year risk and the mean predicted two-year risk compared with the observed two-year risk for each risk group separately, with an overlaid LOWESS smoother. The observed two-year risk for each group was estimated using the Kaplan-Meier survival function to account for censored observations.

The c-statistic and calibration plots were assessed overall and in subgroups of patient data. Subgroups included age (10-year age bands from 40 to 90+ years), co-occurring symptoms (recorded in the three months before baseline FBC), and time span of repeat FBCs (6-monthly time bands from zero to five years). We also assessed the c-statistic and calibration by the number of repeat tests available per patient that were used to define their trend (from two to 10), but as older patients are likely to have more repeat FBCs than younger patients, c-statistics from this analysis may be influenced by increasing age. We therefore stratified this analysis by age group. We compared the c-statistic of the models with that of blood test abnormality (yes/no) on the most recent test in the longitudinal window: low haemoglobin if <13g/dL for men and <11.5g/dL for women, low MCV if <76fL, and raised platelets if >400 x 10^9^/L[22]. The c-statistic for blood test abnormality was derived using a Cox model that included all three tests as binary variables (abnormal=yes/no), with adjustment for age at the test date. We further derived the c-statistic for early-stage (stage I-II) and late-stage (stage III-IV) cancer diagnoses.

Patients were categorised into low-vs high-risk groups using thresholds for two-year risk corresponding to the 1^st^ percentile, every 5^th^ percentile from 5^th^ to 95^th^, and 99^th^ percentile of predicted risks derived in this dataset. Sensitivity, specificity, positive predictive value (PPV), and negative predictive value (NPV) were derived at each threshold.

We derived the two-year cancer incidence using patient data until 2014 to offer a direct comparison with the cancer incidence in the derivation study, which used data over 2000-2014.

### Sensitivity analysis

We performed two sensitivity analyses. First, including cancer diagnoses from CPRD, HES, and ONS databases, in addition to NCRAS. Second, using a traditional cohort study design. Details for both analyses are described in the Supplementary Methods.

## Results

### Summary of patient data

We included 2,956,977 males and 3,561,349 females (Figure 1), with 12,578 (0.4%) and 11,939 (0.3%) diagnosed two years after their baseline FBC, respectively, comparable to patient data over 2000-2014 alone: 0.5% males and 0.4% females. Patients with colorectal cancer were on average around 10 years older than patients without (Table 1). White patients were most likely to get diagnosed (men: 93.0% cancer, 81.8% cancer-free; women: 91.8% cancer, 81.9% cancer-free). Median time (years) from first to last blood test used to derive trends was higher in cases than cancer-free patients: male cases 3.2 and cancer-free 2.7; female cases 3.0 and cancer-free 2.1 (Table S1).

**Table 1:** Summary of patient data.

|  | Males (n=2,956,977) |  | Females (n=3,561,349) |  |
| --- | --- | --- | --- | --- |
|  | Diagnosed | Not diagnosed | Diagnosed | Not diagnosed |
| **Total patients** | 12,578 (0.4%) | 2,944,399 (99.6%) | 11,939 (0.3%) | 3,549,410 (99.7%) |
| **Age (years) at baseline FBC:** |  |  |  |  |
| Mean (SD) | 71.6 (10.3) | 61.6 (13.7) | 73.9 (11.3) | 62.7 (15.0) |
| Median (IQR) | 72 (65-79) | 60 (50-72) | 76 (87-82) | 61 (50-75) |
| **Ethnicity, n (%):** |  |  |  |  |
| Asian | 78 (0.6%) | 46,355 (1.6%) | 75 (0.6%) | 52,873 (1.5%) |
| Black | 203 (1.6%) | 95,943 (3.3%) | 201 (1.7%) | 119,413 (3.4%) |
| Mixed | 28 (0.2%) | 17,342 (0.6%) | 31 (0.3%) | 22,373 (0.6%) |
| Other | 81 (0.6%) | 44,823 (1.5%) | 77 (0.6%) | 50,244 (1.2%) |
| South Asian | 218 (1.7%) | 106,851 (3.6%) | 151 (1.3%) | 117,228 (3.3%) |
| White | 11,691 (93.0%) | 2,407,565 (81.8%) | 10,961 (91.8%) | 2,907,510 (81.9%) |
| Unknown | 279 (2.2%) | 225,520 (7.7%) | 443 (3.7%) | 279,769 (7.9%) |
| **IMD quintile** |  |  |  |  |
| 1 (richest) | 2,835 (21.1%) | 641,060 (21.8%) | 2,776 (21.5%) | 783,492 (22.1%) |
| 2 | 2,833 (21.1%) | 628,975 (21.4%) | 2,719 (21.1%) | 763,896 (21.5%) |
| 3 | 2,736 (20.4%) | 578,092 (19.6%) | 2,595 (20.1%) | 697,553 (19.7%) |
| 4 | 2,565 (19.1%) | 565,080 (19.2%) | 2,459 (19.0%) | 678,795 (19.1%) |
| 5 (poorest) | 2,429 (18.1%) | 525,951 (17.9%) | 2,347 (18.2%) | 619,595 (17.5%) |
| Unknown | 25 (0.2%) | 4,396 (0.2%) | 23 (0.2%) | 5,099 (0.1%) |
| **Symptom, n (%):** |  |  |  |  |
| Any CRC symptom | 782 (6.6%) | 203,777 (6.9%) | 972 (8.3%) | 269,223 (7.6%) |
| Abdominal pain | 273 (2.2%) | 88,848 (3.0%) | 391 (3.2%) | 134,010 (3.8%) |
| Appetite loss | 26 (0.2%) | 3,737 (0.1%) | 20 (0.2%) | 5,597 (0.2%) |
| Change in bowel habit | 62 (0.5%) | 14,162 (0.5%) | 56 (0.5%) | 16,581 (0.5%) |
| Constipation | 152 (1.2%) | 26,515 (0.9%) | 168 (1.4%) | 37,024 (1.0%) |
| Diarrhoea | 136 (1.1%) | 35,511 (1.2%) | 212 (1.8%) | 48,016 (1.4%) |
| Rectal bleeding | 126 (1.0%) | 26,005 (0.9%) | 137 (1.2%) | 21,859 (0.6%) |
| Weight loss | 65 (0.5%) | 19,423 (0.7%) | 74 (0.6%) | 21,164 (0.6%) |
| **Cancer stage, n (%):** |  |  |  |  |
| I | 1,732 (13.8%) |  | 1,348 (11.3%) |  |
| II | 2,904 (23.1%) |  | 2,651 (22.2%) |  |
| III | 2,968 (23.6%) |  | 2,787 (23.3%) |  |
| IV | 2,204 (17.5%) |  | 1,999 (16.7%) |  |
| Unknown | 3,615 (22.0%) |  | 4,134 (26.4%) |  |

Abbreviations: SD=standard deviation; IQR=interquartile range; CRC=colorectal cancer

**Figure 1:**
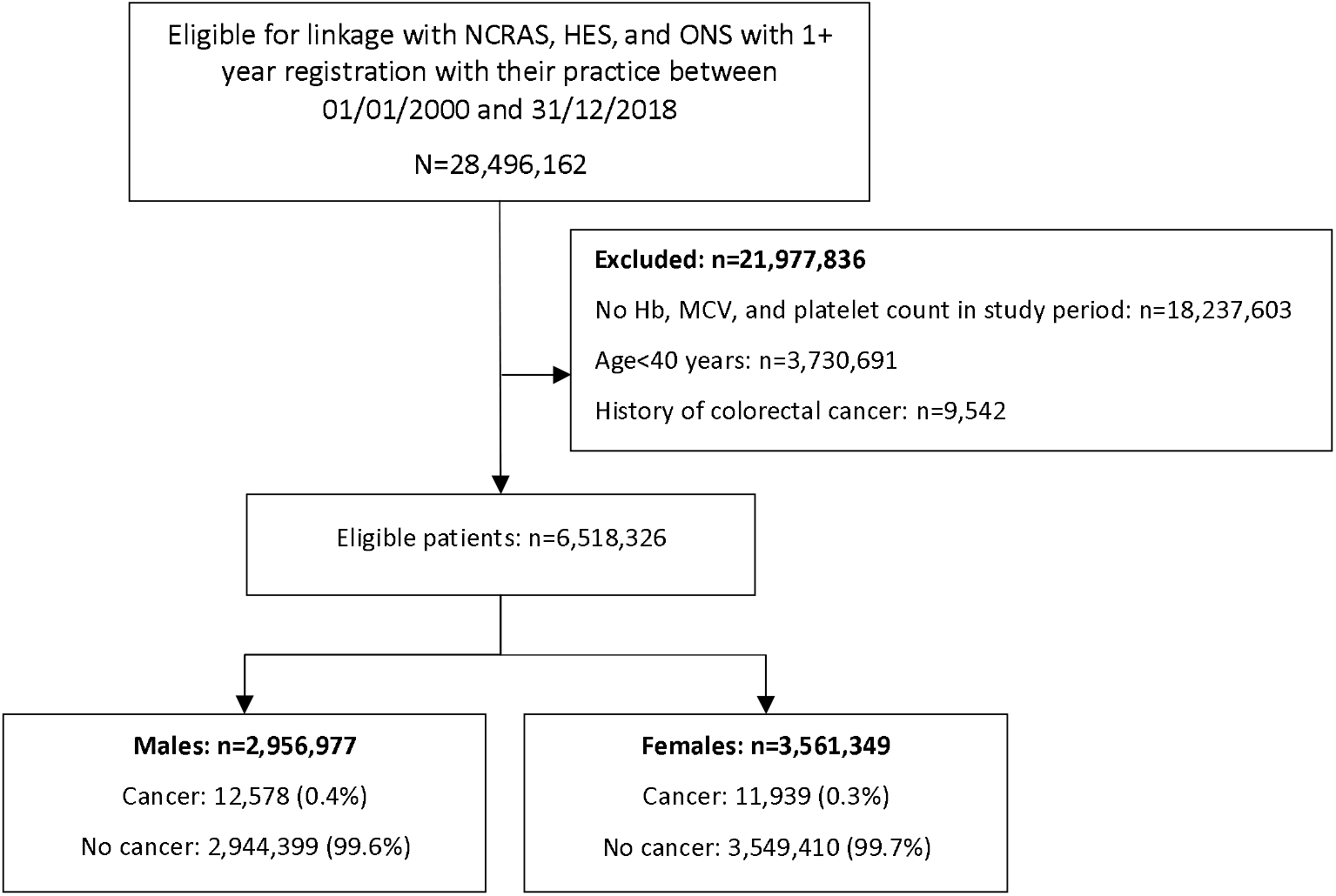
Patient flow diagram. Abbreviations: NCRAS=National Cancer Registration and Analysis Service; HES=Hospital Episode Statistics; ONS=Office of National Statistics; Hb=haemoglobin; MCV=mean corpuscular volume.

### Model performance

The median (range) of predicted risk for men was 0.004 (0.001-0.057) among cases and 0.002 (0.001-0.077) among cancer-free patients. For women, this was 0.003 (0.001-0.049) and 0.001 (0.001-0.099), respectively. The c-statistic (95% CI) was 0.73 (0.72-0.73) for males and 0.74 (0.74-0.75) for females (Table 2). The calibration slope (95% CI) was 0.92 (0.89-0.94) for males and 0.95 (0.93-0.98) for females. Calibration plots showed increasing under-prediction with increasing and was largest for the highest risk group (Figure 2): 0.14% predicted vs 0.16% observed risk in males and 0.10% predicted vs 0.12% observed risk in females. In males, the c-statistic (95% CI) was 0.72 (0.72-0.73) for early-stage diagnoses and 0.71 (0.71-0.72) for late-stage. In females, it was 0.74 (0.73-0.74) for early-stage diagnoses and 0.72 (0.71-0.72) for late-stage.

**Table 2:** Performance measures (95% CI) of the BLOODTRACC models.

| Performance measure | Males | Females |
| --- | --- | --- |
| Brier score | 0.0042 | 0.0033 |
| C-statistic | 0.73 (0.72 - 0.73) | 0.74 (0.74 - 0.75) |
| D-statistic | 1.18 (1.16 - 1.21) | 1.32 (1.29 - 1.35) |
| Calibration slope | 0.92 (0.89-0.94) | 0.95 (0.93-0.98) |

**Figure 2:**
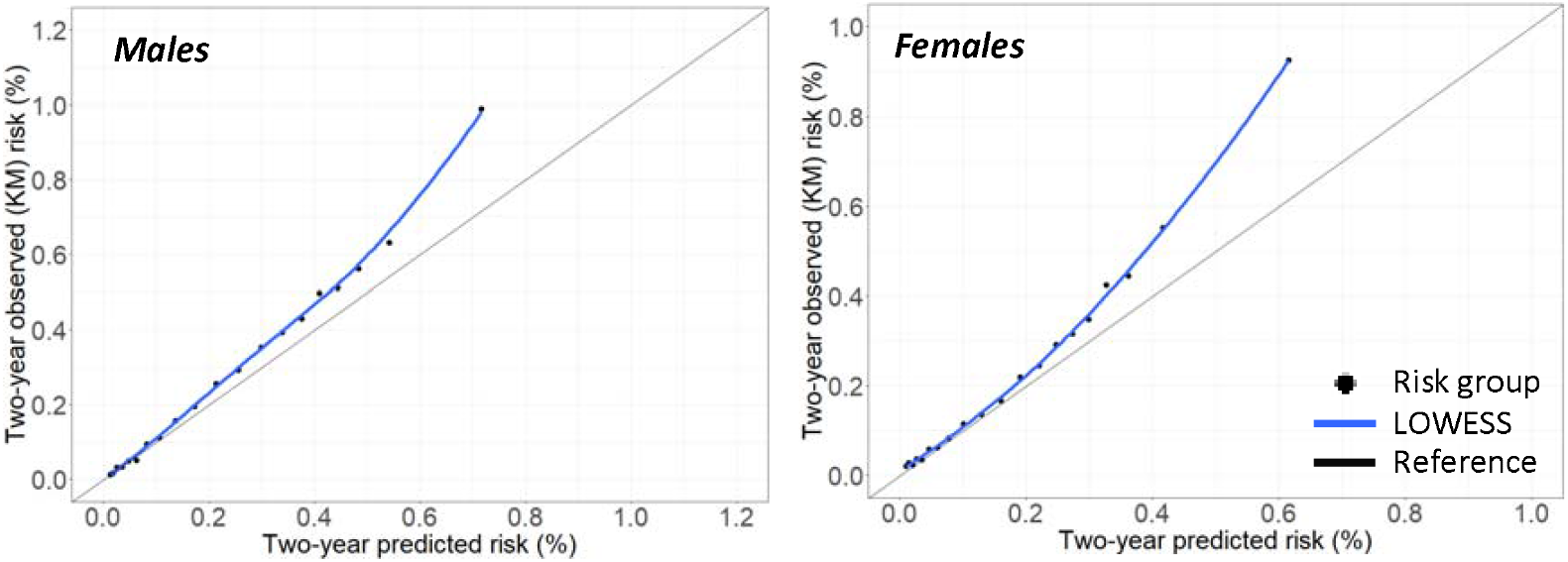
Calibration plots for the BLOODTRACC models. Abbreviations: KM=Kaplan-Meier

### Model performance in patient subgroups

The c-statistic generally decreased as males and females grew older, ranging 0.54-0.63 and 0.58-0.63, respectively (Figure S1). Calibration plots showed good calibration in patients aged <50 years, with increasing under-prediction with increasing age, with most under-prediction observed for patients aged 80-89 years and greater for men than women (Figure S2). The models slightly over-predicted risk in each ethnic group, but under-estimated in White patients and higher IMD quintiles. The c-statistic (95% CI) across ethnic groups ranged 0.72-0.79 for men and 0.68-0.81 for women (Figure S3), lowest for White men (0.72 (0.72-0.72)) and South Asian women (0.68 (0.64-0.72)), and similar in IMD quintile groups (Figure S4). The c-statistic (95% CI) was higher in men without constipation (with 0.66 (0.62-0.70); without 0.75 (0.74-0.75)) or with rectal bleeding (with 0.79 (0.76-0.83); without (0.75 (0.74-0.75)) and women without appetite loss (with 0.62 (0.52-0.73); without (0.74 (0.74-0.74)), constipation (with 0.66 (0.62-0.69); without (0.74 (0.74-0.74)), or diarrhoea (with 0.69 (0.66-0.72); without (0.74 (0.74-0.74)) (Figure S5). The symptom subgroups that had the highest c-statistics were rectal bleeding and abdominal pain for both men and women. For each age group, the c-statistic in subgroups formed by the number of repeat tests available per patient that were used to derive trends ranged 0.56-0.72 if aged 40-49 years and 0.50-0.70 if aged 90+ years in men and 0.53-0.73 if aged 40-49 years and 0.56-0.65 if aged 90+ years in women (Figure S6 and Figure S7).

When we assessed the c-statistic by the time span encompassing the all repeat tests, the c-statistic was similar for all time spans up to 2.0-2.5 years (range 0.74-0.76) for men and 2.5-3.0 years (range 0.72-0.77) for women, but decreased as the time span of repeat tests grew larger thereafter: 0.73 (95% CI=0.71-0.74) for repeat tests over 2.5-3.0 years to 0.68 (95% CI=0.67-0.69) over 4.5-5.0 years in men and 0.73 (95% CI=0.72-0.75) for repeat tests over 3.0-3.5 years to 0.70 (95% CI=0.69-0.71) over 4.5-5.0 years in women (Figure S8). The models were well calibrated among patients with up to three repeat tests, with increasing under-prediction as the number of repeat tests increased, and among patients with all their repeat tests confined within a six-month period, with under-prediction observed for longer repeat testing periods (Figure S9).

### Comparison to abnormal test results

For each age group, the c-statistic was comparable between the BLOODTRACC models, which included blood test trends, and Cox models, which included the abnormality counterparts (Figure S7). However, the c-statistic for the BLOODTRACC models appeared to become greater than for Cox models including abnormality in women aged at least 70 years.

### Diagnostic accuracy

The highest risk percentile, 99%, corresponded to a risk threshold of 0.7810% for males and 0.6872% for females, with 3.56% and 5.03% sensitivity and 99.01% and 99.01% specificity, respectively (Table S2 and Table S3). The PPV increased as the threshold used to define low-vs high-risk increased, ranging 0.43-1.52% for men and 0.34-1.68% for women. The NPV ranged 99.59-99.98% for men and 99.68-99.96% for women, indicating a high proportion of patients with low predicted risk without an observed diagnosis regardless of the threshold used to define low-vs high-risk, likely due to the low event rate overall.

### Sensitivity analysis

In the first sensitivity analysis, including cancer diagnoses from NCRAS, CPRD, HES, and ONS increased the c-statistic (95% CI) increased to 0.747 (0.744-0.750) for males and remained similar at 0.740 (0.736-0.744) for females, with an increased event rate and therefore increased under-prediction (Table S4, Figure S10). In the second sensitivity analysis, a traditional cohort study design increased the c-statistic (95% CI) to 0.84 (0.83-0.85) in men and 0.91 (0.89-0.93) in women, with comparable event rates (Table S5). Further results are in the Supplementary Results.

## Discussion

### Summary of main findings

Our prediction models have good discriminative ability for two-year risk of colorectal cancer, based on only age, sex, and trends in haemoglobin, MCV, and platelet count earlier than two years before diagnosis, and performed slightly better for early-stage diagnoses than late-stage. The models however under-predicted two-year risk in the patients with higher predicted risks, likely reflecting older patients, as calibration in younger patients was good. The two-year event rate remained the same when considering only patient data over 2000-2014, coinciding with the prior derivation study period, and was comparable to the derivation study (0.4% in men, 0.3% in women)[16]. This under-prediction may be explained by practice-level differences between practices contributing to the CPRD GOLD and AURUM databases.

### Strengths and limitations

A key strength of this study was the large sample size and follow-up duration used to validate the prediction models. This allowed us to explore the impact of patient age and varying time periods and number of tests capturing a trend on model performance. One limitation is that the reason for the FBC being ordered in primary care is unknown. The FBC is a non-specific test, so are ordered in primary care for many reasons and not specifically for colorectal cancer. Knowing the reason for testing could help identify other conditions that could influence blood test trends. Additionally, it is possible that patients without colorectal cancer who have many FBCs in the five-year period have another disease or condition that influences blood levels over time. Therefore, some false positives (patients determined to be high risk who are not diagnosed with colorectal cancer) may have another illness. Data on comorbidities, including other cancers, will be accounted for as future work.

### Comparison with existing literature

Our systematic review identified 13 prediction models that use some FBC data to inform risk of colorectal cancer[23]. All of these models are static, using a single test from one time point per patient, except the ColonFlag model, a machine-learning algorithm, derived using data from an Israeli population, designed to predict three-to-six month colorectal cancer risk based on changes in all FBC components measured at 36 and 18 months before a patient’s current FBC[24]. The use of repeated measures data can provide more individualised risk predictions, so our models may offer improved risk stratification to existing static models (using covariates measured at one time point). However, predictive performance of our dynamic BLOODTRACC models is yet to be compared to that of static models in the same patient cohort using similar study designs to reduce heterogeneity.

We previously reported an external validation of the ColonFlag, performed in the same split sample internal validation cohort used in the BLOODTRACC model derivation study to offer a direct comparison of model performance[16]. The c-statistic for two-year risk was comparable between our models and the ColonFlag. In men, it was 0.75 for BLOODTRAC in this external validation, 0.75 for BLOODTRACC in its derivation study, and 0.76 for ColonFlag in its previous external validation. For women, it was 0.74 for BLOODTRAC in this external validation, 0.76 for BLOODTRACC in its derivation study, and 0.76 for ColonFlag in its previous external validation. Performance of each model was also similar in subgroups of age, number of FBCs used to derive trend, and length of the longitudinal window. Comparable performance was expected, as the models use the same data (age, sex, changes over time in FBC tests) to identify risk, although the ColonFlag includes trends in all 20 FBC parameters to identify risk[24] and our models are simpler, using only haemoglobin, MCV, and platelets. Discrimination remained comparable regardless, suggesting these additional FBC parameters may not enhance risk estimation. The ColonFlag model is commercially developed and not publicly available. The BLOODTRACC models may increase the likelihood of adoption and embedding within electronic health record systems, facilitate flagging of cancer risk in practice.

Our second systematic review, which focused on clinical prediction models incorporating trends over repeated blood tests (liver function, renal function, and FBC) to inform cancer diagnosis, did not identify any additional existing trend-based prediction models for colorectal cancer, other than our current BLOODTRACC models[16] and the ColonFlag algorithm[24].

### Implications for practice

Our dynamic prediction models are designed to provide an up-to-date risk prediction each time a new haemoglobin, MCV, or platelet measurement is added to a patient’s record. Using combinations of trends over repeated tests could identify subtle simultaneous cancer-relevant changes in tests that could otherwise be missed in practice, including changes within the normal reference range, which are unlikely to be noticed by a clinician. The models are designed to use routinely available data, accounting for the sporadic and irregular nature of blood testing in primary care, and we plan for them to be programmed into practice software to run automatically when a new FBC becomes available. Therefore, there will likely be minimal additional work for patients or GP staff to identify a patient’s risk of undiagnosed colorectal cancer.

FIT testing, which examines stool samples for traces of blood, has proved a useful test outside the screening programme for ruling out colorectal cancer in patients with symptoms attending primary care, based on a 98% NPV in those with a negative FIT in a recent primary care study[25, 26]. Patients identified as high-risk from our models could be offered a FIT test, which is much more practical, cheaper and less invasive than colonoscopy. As the use of FIT increases in clinical practice and FIT results are recorded in the primary care electronic health record, research should investigate the additional diagnostic value of adding historical blood test trend to the FIT value at the time of clinical presentation. This would complement past and ongoing efforts to increase the predictive value of FIT by combining it in a model with patient characteristics and blood tests taken at the time of the FIT[27–30]. Further work could also aim to modify the FIT threshold considered ‘positive’ in high-risk patients identified from the BLOODTRACC models.

Prediction models should be updated regularly to incorporate changes in clinical practice[31]. The models have good discriminative ability using only data measured earlier than two years before diagnosis, but discrimination could be improved by updating the model to include data closer to diagnosis, enhancing risk stratification. For example, our sensitivity analysis using a cohort study design in essence extended the trend to capture changes measured closer to diagnosis, which improved the c-statistic. This approach would introduce bias in that cancer cases have more frequent testing than cancer-free patients in routinely collected data. However, the updated model could include the number of repeat tests used to derive each patient’s trend as a covariate to adjust for this imbalance. As model discrimination was slightly better for younger individuals, BLOODTRAC may have an important role in enhancing the detection of early-onset colorectal cancer, which is rising in incidence[32]. Accounting for comorbidity in the model may improve discrimination in older patients, where blood test abnormalities are less likely to be caused by cancer than in younger less comorbid patients. These updates could also reduce the degree of under-prediction identified in this study.

### Conclusion

Our dynamic clinical risk prediction models perform well in external primary care data, relying only on data measured earlier than two years before diagnosis, but did not outperform blood test abnormality, recommended in referral guidelines to inform referral. Extending the trend closer to diagnosis may improve predictive performance.

## Additional information

## Acknowledgements

The authors would also like to thank Patient and Public Involvement contributors Sue Dutton, Alton Sutton, Bernard Gudgin, Clara Martins de Barros, Emily Lam, Ian Blelloch, Julian Ashton, Margaret Ogden, Shannon Draisey, and Susan Lynne for applying a patient perspective on the use of blood tests to improved cancer detection and the acceptability of repeat testing in primary care.

## Funding

This work was funded by a National Institute for Health and Care Research (NIHR) School for Primary Care Research (SPCR) Post-doctoral fellowship for this work (award number: C092) and the NIHR Policy Research Programme (Policy Research Unit on Cancer Awareness, Screening and Early Diagnosis, reference PR-PRU-NIHR206132). This report presents independent research and the views expressed are those of the authors and not necessarily those of the NIHR, SPCR, or Department of Health and Social Care.

## Competing interests

The authors declare no competing interests.

## Data availability

The dataset used is available from the authors but is subject to access approval by the CPRD[33].

## Notes

### Competing Interest Statement

The authors have declared no competing interest.

### Author Declarations

Research Data Governance Committee of the Clinical Practice Research Datalink gave ethical approval for this work

