## Supplementary file for "Full BLOOD count TRends for colorectal cAnCer deteCtion (BLOODTRACC): external validation of colorectal cancer prediction models in English primary care"

**Supplementary Methods – Sensitivity Analysis**

The main analysis uses cancers from NCRAS to offer a direct comparison to the performance measures derived in the derivation study, as only cancers from the NCRAS database were used for model derivation. In our CPRD AURUM external validation dataset, cancers were also available from CPRD, HES, and ONS databases. We therefore performed a sensitivity analysis to include diagnoses from these additional sources. We first re-derived the validation cohort, excluding patients with a history of cancer from NCRAS, CPRD, HES, or ONS. The outcome was defined as before, except diagnoses were primarily selected from the NCRAS database and additional diagnoses were taken as the earliest diagnosis in CPRD, HES, or ONS if not identified in NCRAS.

We performed a second sensitivity analysis using a different study design. For the main analysis, we performed a hybrid case-control and cohort study that excluded haemoglobin, MCV, and platelet measurements within the two years prior to diagnosis in cases and study exit in cancer-free patients, following the design of the model derivation study. This approach ensures each patient has at least two years of follow-up, but does not entirely reflect patient flow in practice, as patients can be diagnosed within two years of their FBC test. This sensitivity analysis therefore used a traditional cohort study design. For each patient, we identified the current/latest FBC, which was considered the baseline test, and trends were identified using all historical tests available up to five years prior to the baseline test. This sensitivity analysis did not include a two-year washout period prior to diagnosis in cases and study exit in cancer-free patients. The outcome was therefore a diagnosis of colorectal cancer within two years following the baseline test. Patients without a diagnosis in this two-year period were censored at the earliest of date of leaving the practice, death, 31st December 2018, or two years after their baseline test. All other study details remained the same.

**Supplementary Results – Sensitivity Analysis**

Including cancer diagnoses from NCRAS, CPRD, HES, and ONS resulted in the inclusion of 2,956,977 men and 3,561,349 women, with the number with a cancer diagnosis increasing from 12,578 (0.4%) and 11,939 (0.3%) to 13,423 (0.5%) and 12,919 (0.4%), respectively (Table S4). The c-statistic (95% CI) increased slightly to 0.747 (0.744-0.750) for males and remained similar at 0.740 (0.736-0.744) for females. The calibration slope (95% CI) was 0.91 (0.88-0.93) for males and 0.94 (0.92-0.97) for females. Calibration plots indicated that the models consistently under-predicted risk, with under-prediction worsening from the main analysis, as expected due to the inclusion of further observed diagnoses. Under-prediction was greatest for the highest risk group (Figure S10): 0.69% predicted vs 1.28% observed risk in males and 0.62% predicted vs 1.00% observed risk in females.

The second sensitivity analysis using a cohort study design included 5,011,297 men and 7,010,202 women, of which 0.4% (n=18,876) and 0.3% (n=17,501) were diagnosed with colorectal cancer within two years following their baseline blood test, respectively (Table S5). This was comparable to the main analysis using a hybrid case-control and cohort study design: 0.4% in men and 0.3% in women. This approach, which Included data measured closer to diagnosis, increased the c-statistic (95% CI) from 0.73 (0.72-0.73) to 0.84 (0.83-0.85) in men and from 0.74 (0.74-0.75) to 0.91 (0.89-0.93) in women.

### Supplementary Tables and Figures

#### Table S1: Summary of haemoglobin, MCV, and platelet data and follow-up time

| | Males | | Females | |
|---|---|---|---|---|
| | **Diagnosed** | **Not diagnosed** | **Diagnosed** | **Not diagnosed** |
| **No. tests in total** | 11,985 | 3,557,891 | 12,614 | 2,951,603 |
| **Mean (SD) no. tests** | 5.9 (7.2) | 4.2 (5.0) | 5.3 (6.2) | 3.7 (4.4) |
| **Median time (range)[1]** | 3.2 (0-5.0) | 2.7 (0-5.0) | 3.0 (0-5.0) | 2.1 (0-5.0) |
| **Median follow-up (range)[2]** | 5.18 (1.75-7.24) | 4.69 (1.75-7.25) | 5.00 (1.75-7.25) | 4.11 (1.75-7.25) |

[1]Time (years) between the first and last FBC in the five-year longitudinal period.
[2]Time (years) from first FBC to diagnosis/censor date.
Abbreviations: MCV=mean corpuscular volume; SD=standard deviation

**Figure S1: C-statistic for the BLOODTRACC models by age (years) at baseline in males (top) and females (bottom)**

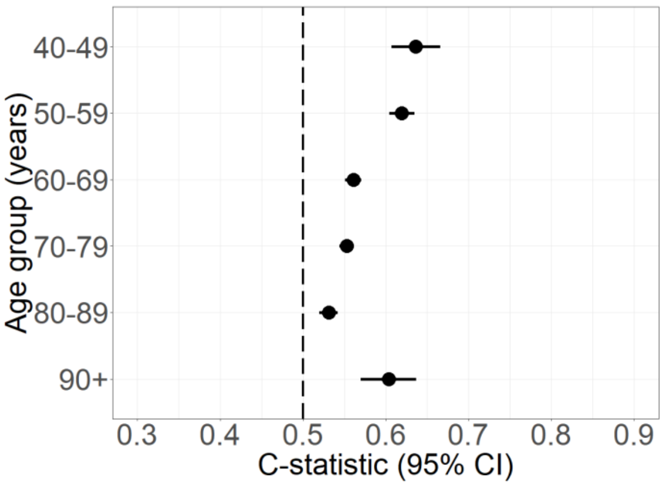

| Age group (years) | No. patients | No. (%) cases | C-statistic (95% CI) |
|---|---|---|---|
| 40-49 | 697362 | 328 (0.05%) | 0.64 (0.61-0.67) |
| 50-59 | 710610 | 1265 (0.18%) | 0.62 (0.60-0.63) |
| 60-69 | 651765 | 3357 (0.52%) | 0.56 (0.55-0.57) |
| 70-79 | 530148 | 4628 (0.87%) | 0.55 (0.54-0.56) |
| 80-89 | 318943 | 2723 (0.85%) | 0.53 (0.52-0.54) |
| 90+ | 48149 | 277 (0.58%) | 0.60 (0.57-0.64) |

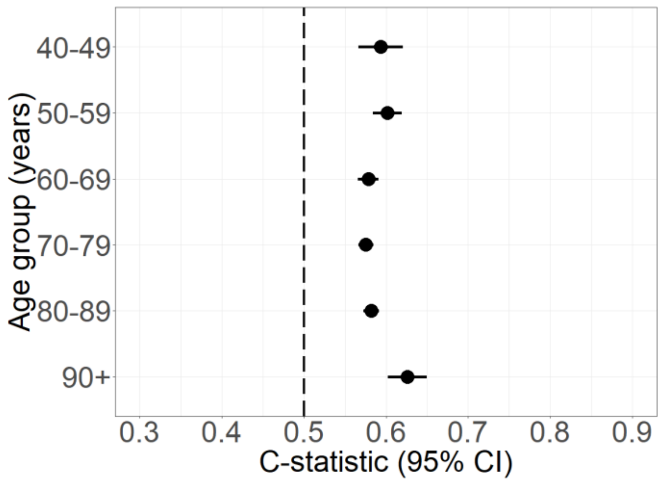

| Age group (years) | No. patients | No. (%) cases | C-statistic (95% CI) |
|---|---|---|---|
| 40-49 | 877079 | 446 (0.05%) | 0.59 (0.57-0.62) |
| 50-59 | 795676 | 1018 (0.13%) | 0.60 (0.58-0.62) |
| 60-69 | 666607 | 2191 (0.33%) | 0.58 (0.57-0.59) |
| 70-79 | 595458 | 4006 (0.67%) | 0.58 (0.57-0.58) |
| 80-89 | 493631 | 3704 (0.75%) | 0.58 (0.57-0.59) |
| 90+ | 132898 | 574 (0.43%) | 0.63 (0.60-0.65) |

**Figure S2: Calibration plots for the BLOODTRACC models by age (years) at baseline, ethnicity, and IMD quintile in males (left) and females (right)**

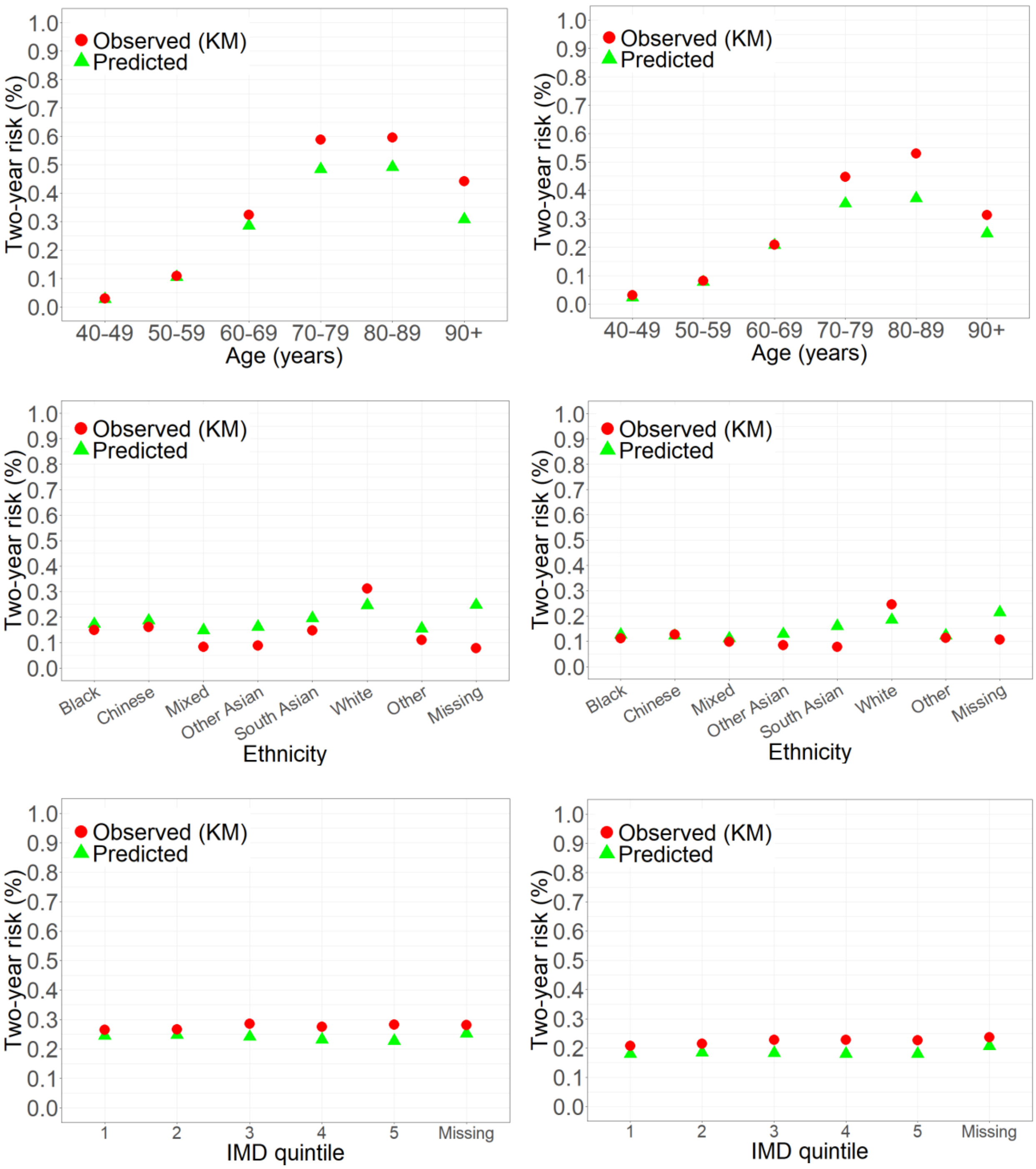

Abbreviations: KM=Kaplan-Meier

**Figure S3: C-statistic for the BLOODTRACC models by ethnicity in males (top) and females (bottom)**

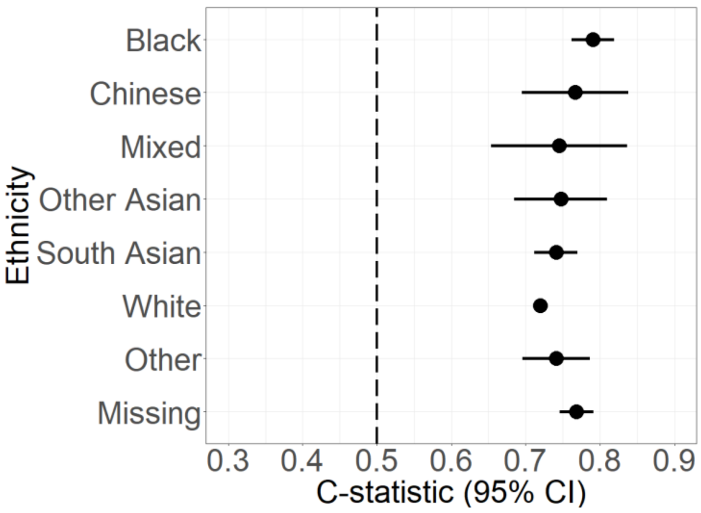

| No. patients | No. (%) cases | C-statistic (95% CI) |
|---|---|---|
| 96146 | 203 (0.21%) | 0.79 (0.76-0.82) |
| 9404 | 30 (0.32%) | 0.77 (0.70-0.84) |
| 17370 | 28 (0.16%) | 0.75 (0.65-0.84) |
| 37029 | 48 (0.13%) | 0.75 (0.68-0.81) |
| 107069 | 218 (0.20%) | 0.74 (0.71-0.77) |
| 2419256 | 11691 (0.48%) | 0.72 (0.72-0.72) |
| 44904 | 81 (0.18%) | 0.74 (0.70-0.79) |
| 225799 | 279 (0.12%) | 0.77 (0.75-0.79) |

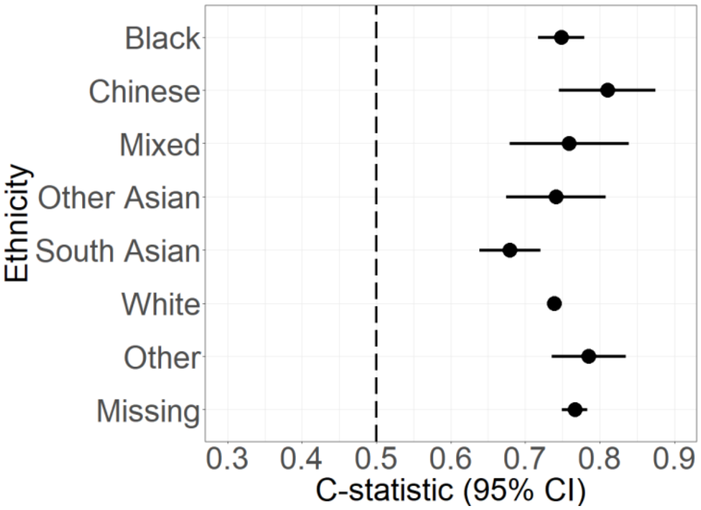

| No. patients | No. (%) cases | C-statistic (95% CI) |
|---|---|---|
| 119614 | 201 (0.17%) | 0.75 (0.72-0.78) |
| 13967 | 28 (0.2%) | 0.81 (0.75-0.88) |
| 22404 | 31 (0.14%) | 0.76 (0.68-0.84) |
| 38981 | 47 (0.12%) | 0.74 (0.67-0.81) |
| 117379 | 151 (0.13%) | 0.68 (0.64-0.72) |
| 2918471 | 10961 (0.38%) | 0.74 (0.73-0.74) |
| 50321 | 77 (0.15%) | 0.79 (0.74-0.83) |
| 280212 | 443 (0.16%) | 0.77 (0.75-0.78) |

**Figure S4: C-statistic for the BLOODTRACC models by IMD quintile in males (top) and females (bottom)**

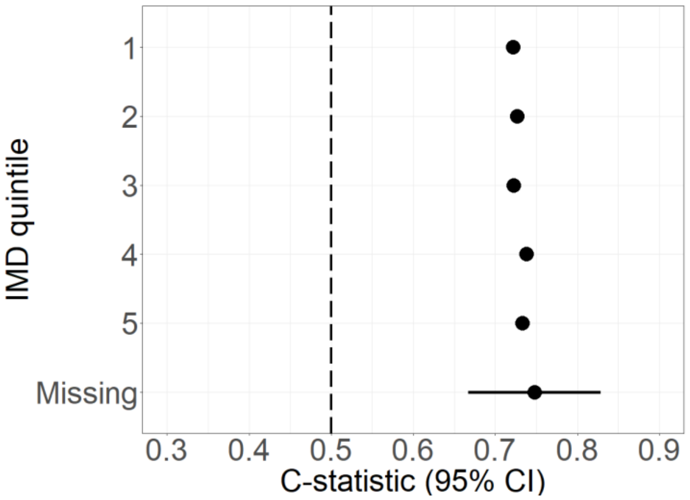

| No. patients | No. (%) cases | C-statistic (95% CI) |
|---|---|---|
| 643895 | 2666 (0.41%) | 0.72 (0.71-0.73) |
| 631808 | 2657 (0.42%) | 0.73 (0.72-0.73) |
| 580828 | 2558 (0.44%) | 0.72 (0.71-0.73) |
| 567645 | 2411 (0.42%) | 0.74 (0.73-0.75) |
| 528380 | 2266 (0.43%) | 0.73 (0.72-0.74) |
| 4421 | 20 (0.45%) | 0.75 (0.67-0.83) |

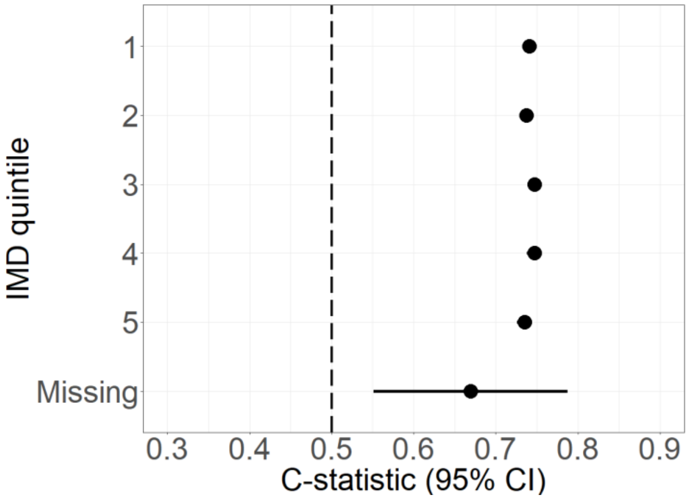

| No. patients | No. (%) cases | C-statistic (95% CI) |
|---|---|---|
| 786268 | 2565 (0.33%) | 0.74 (0.73-0.75) |
| 766615 | 2511 (0.33%) | 0.74 (0.73-0.75) |
| 700148 | 2401 (0.34%) | 0.75 (0.74-0.76) |
| 681254 | 2265 (0.33%) | 0.75 (0.74-0.76) |
| 621942 | 2183 (0.35%) | 0.73 (0.73-0.74) |
| 5122 | 14 (0.27%) | 0.67 (0.55-0.79) |

**Figure S5: C-statistic for the BLOODTRACC models by presence of co-occurring symptoms[1] in males (top) and females (bottom)**

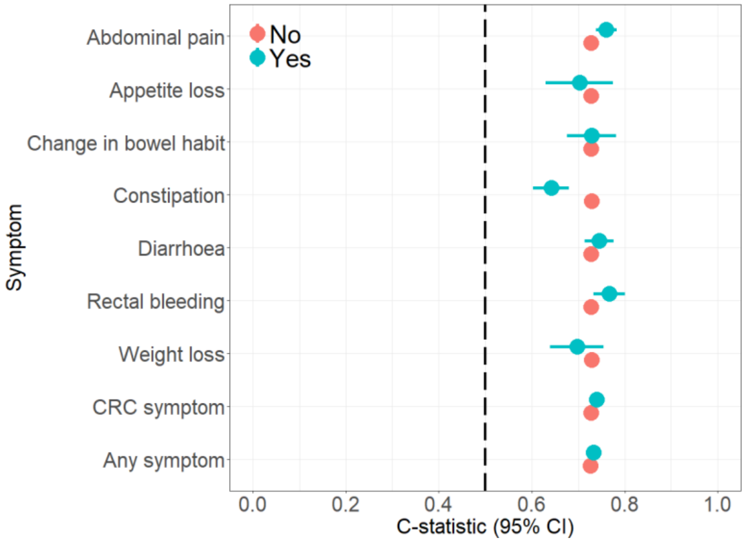

| Group | No. patients | No. (%) cases | C-statistic (95% CI) |
|---|---|---|---|
| Yes | 2867856 | 12305 (0.43%) | 0.73 (0.72-0.73) |
| No | 89121 | 273 (0.31%) | 0.76 (0.74-0.78) |
| Yes | 2953214 | 12552 (0.43%) | 0.73 (0.72-0.73) |
| No | 3763 | 26 (0.69%) | 0.70 (0.63-0.77) |
| Yes | 2942753 | 12516 (0.43%) | 0.73 (0.72-0.73) |
| No | 14224 | 62 (0.44%) | 0.73 (0.68-0.78) |
| Yes | 2930310 | 12426 (0.42%) | 0.73 (0.73-0.73) |
| No | 26667 | 152 (0.57%) | 0.64 (0.60-0.68) |
| Yes | 2921330 | 12442 (0.43%) | 0.73 (0.72-0.73) |
| No | 35647 | 136 (0.38%) | 0.75 (0.71-0.78) |
| Yes | 2930846 | 12452 (0.42%) | 0.73 (0.72-0.73) |
| No | 26131 | 126 (0.48%) | 0.77 (0.73-0.80) |
| Yes | 2937489 | 12513 (0.43%) | 0.73 (0.72-0.73) |
| No | 19488 | 65 (0.33%) | 0.70 (0.64-0.75) |
| Yes | 2752418 | 11796 (0.43%) | 0.73 (0.72-0.73) |
| No | 204559 | 782 (0.38%) | 0.74 (0.73-0.75) |
| Yes | 1943857 | 8469 (0.44%) | 0.73 (0.72-0.73) |
| No | 1013120 | 4109 (0.41%) | 0.73 (0.73-0.74) |

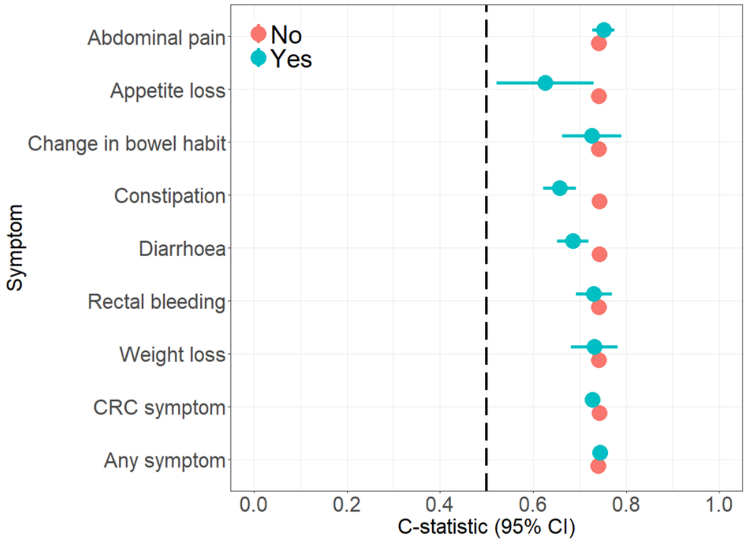

| Group | No. patients | No. (%) cases | C-statistic (95% CI) |
|---|---|---|---|
| Yes | 3426948 | 11548 (0.34%) | 0.74 (0.74-0.74) |
| No | 134401 | 391 (0.29%) | 0.75 (0.73-0.78) |
| Yes | 3555732 | 11919 (0.34%) | 0.74 (0.74-0.75) |
| No | 5617 | 20 (0.36%) | 0.63 (0.52-0.73) |
| Yes | 3544712 | 11883 (0.34%) | 0.74 (0.74-0.75) |
| No | 16637 | 56 (0.34%) | 0.73 (0.66-0.79) |
| Yes | 3524157 | 11771 (0.33%) | 0.74 (0.74-0.75) |
| No | 37192 | 168 (0.45%) | 0.66 (0.62-0.69) |
| Yes | 3513121 | 11727 (0.33%) | 0.74 (0.74-0.75) |
| No | 48228 | 212 (0.44%) | 0.68 (0.65-0.72) |
| Yes | 3539353 | 11802 (0.33%) | 0.74 (0.74-0.75) |
| No | 21996 | 137 (0.62%) | 0.73 (0.69-0.77) |
| Yes | 3540111 | 11865 (0.34%) | 0.74 (0.74-0.75) |
| No | 21238 | 74 (0.35%) | 0.73 (0.68-0.78) |
| Yes | 3291153 | 10966 (0.33%) | 0.74 (0.74-0.75) |
| No | 270196 | 973 (0.36%) | 0.73 (0.71-0.74) |
| Yes | 2286188 | 7550 (0.33%) | 0.74 (0.74-0.74) |
| No | 1275161 | 4389 (0.34%) | 0.74 (0.74-0.75) |

[1]Presence (=yes) or no presence (=no) of the symptom within the three months prior to the current (baseline) test.

**Figure S6: C-statistic for the BLOODTRACC models by number of FBCs per age group in males**

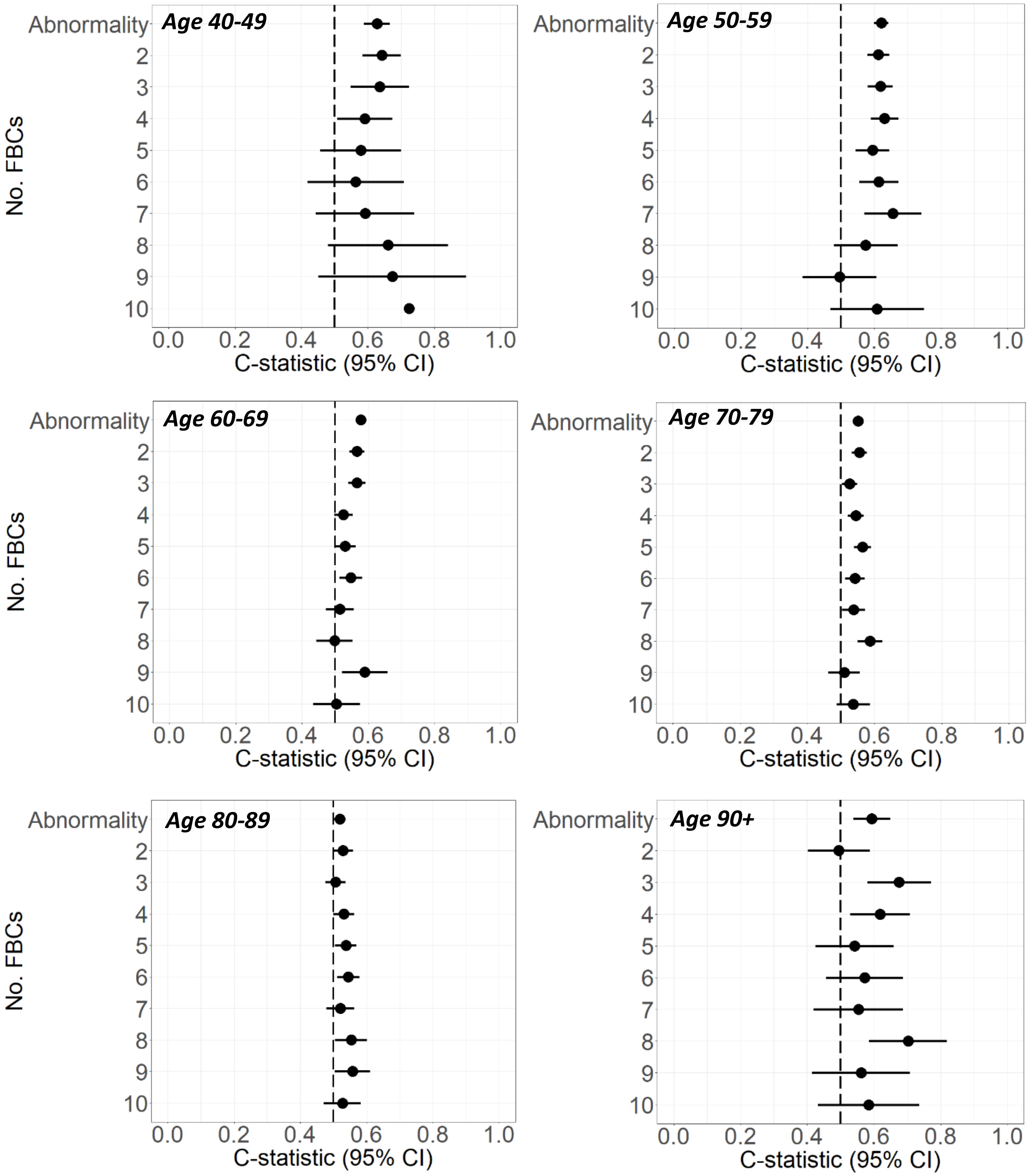

Abbreviations: FBC=full blood count.

**Figure S7: C-statistic for the BLOODTRACC models by number of FBCs per age group in females**

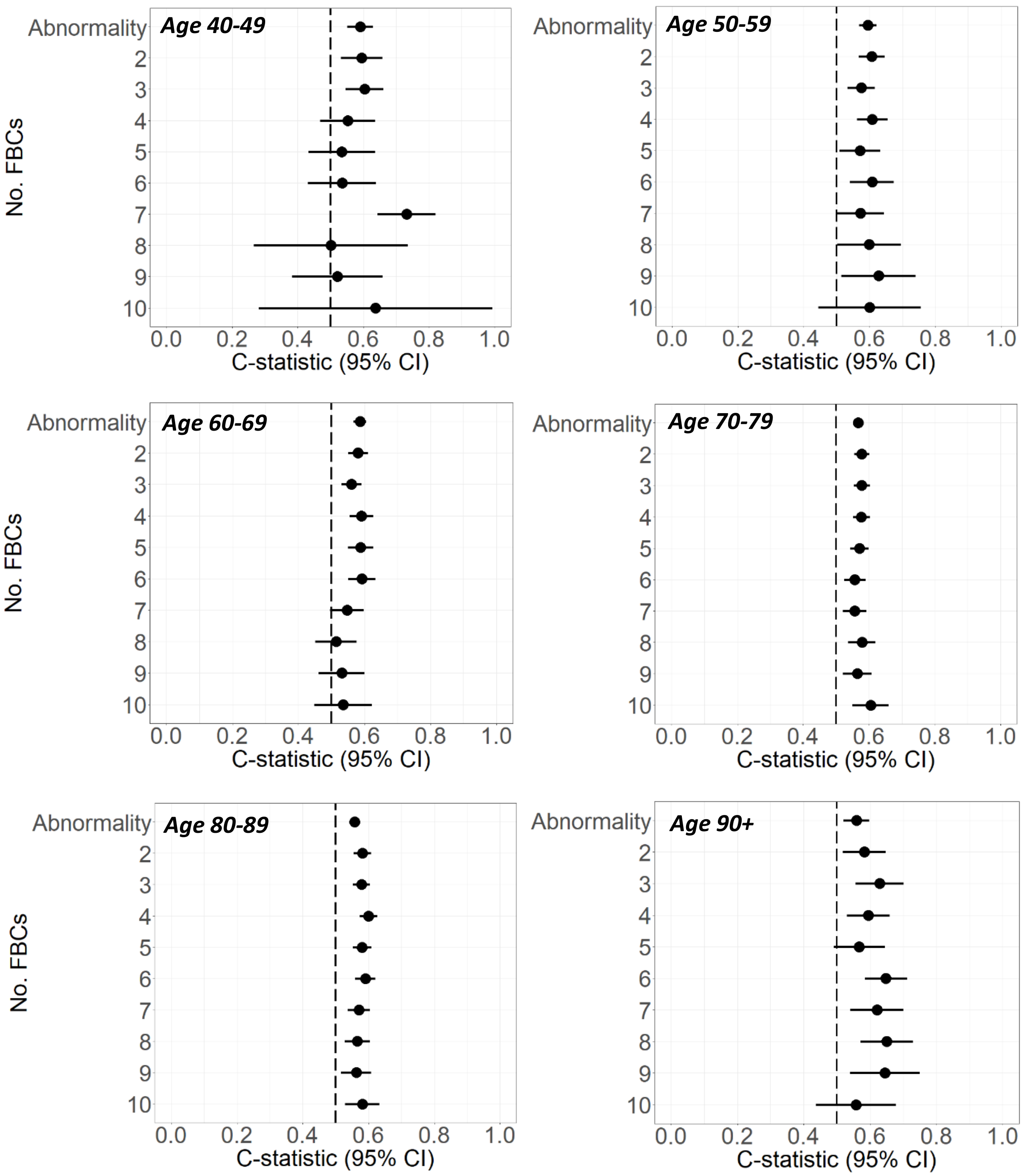

Abbreviations: FBC=full blood count.

**Figure S8: C-statistic for the BLOODTRACC models by time span of FBCs in males (top) and females (bottom)**

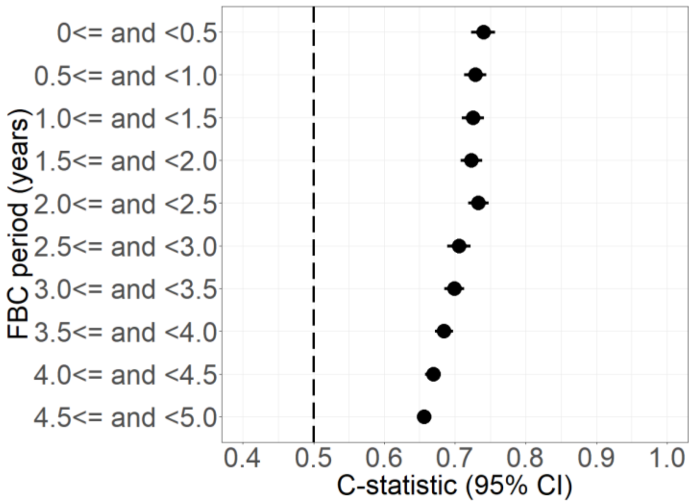

| No. patients | No. (%) cases | C-statistic (95% CI) |
|---|---|---|
| 133949 | 588 (0.44%) | 0.74 (0.72-0.76) |
| 125203 | 617 (0.49%) | 0.73 (0.71-0.74) |
| 143081 | 681 (0.48%) | 0.73 (0.71-0.74) |
| 142994 | 731 (0.51%) | 0.72 (0.71-0.74) |
| 155537 | 747 (0.48%) | 0.73 (0.72-0.75) |
| 164434 | 751 (0.46%) | 0.71 (0.69-0.72) |
| 189532 | 933 (0.49%) | 0.70 (0.68-0.71) |
| 228538 | 1172 (0.51%) | 0.68 (0.67-0.70) |
| 328513 | 1674 (0.51%) | 0.67 (0.66-0.68) |
| 443649 | 2531 (0.57%) | 0.66 (0.65-0.67) |

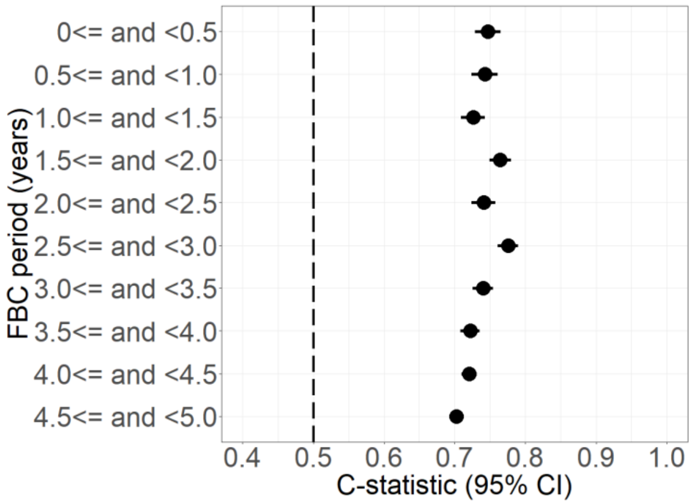

| No. patients | No. (%) cases | C-statistic (95% CI) |
|---|---|---|
| 157361 | 565 (0.36%) | 0.75 (0.73-0.77) |
| 153369 | 541 (0.35%) | 0.74 (0.72-0.76) |
| 171410 | 660 (0.39%) | 0.73 (0.71-0.74) |
| 175031 | 659 (0.38%) | 0.76 (0.75-0.78) |
| 192937 | 678 (0.35%) | 0.74 (0.72-0.76) |
| 210673 | 753 (0.36%) | 0.78 (0.76-0.79) |
| 247959 | 929 (0.37%) | 0.74 (0.73-0.75) |
| 308895 | 1076 (0.35%) | 0.72 (0.71-0.74) |
| 444054 | 1736 (0.39%) | 0.72 (0.71-0.73) |
| 640908 | 2554 (0.4%) | 0.70 (0.69-0.71) |

Abbreviations: FBC=full blood count.

**Figure S9: Calibration plots for the BLOODTRACC models by number of repeat tests and testing period in males (left) and females (right)**

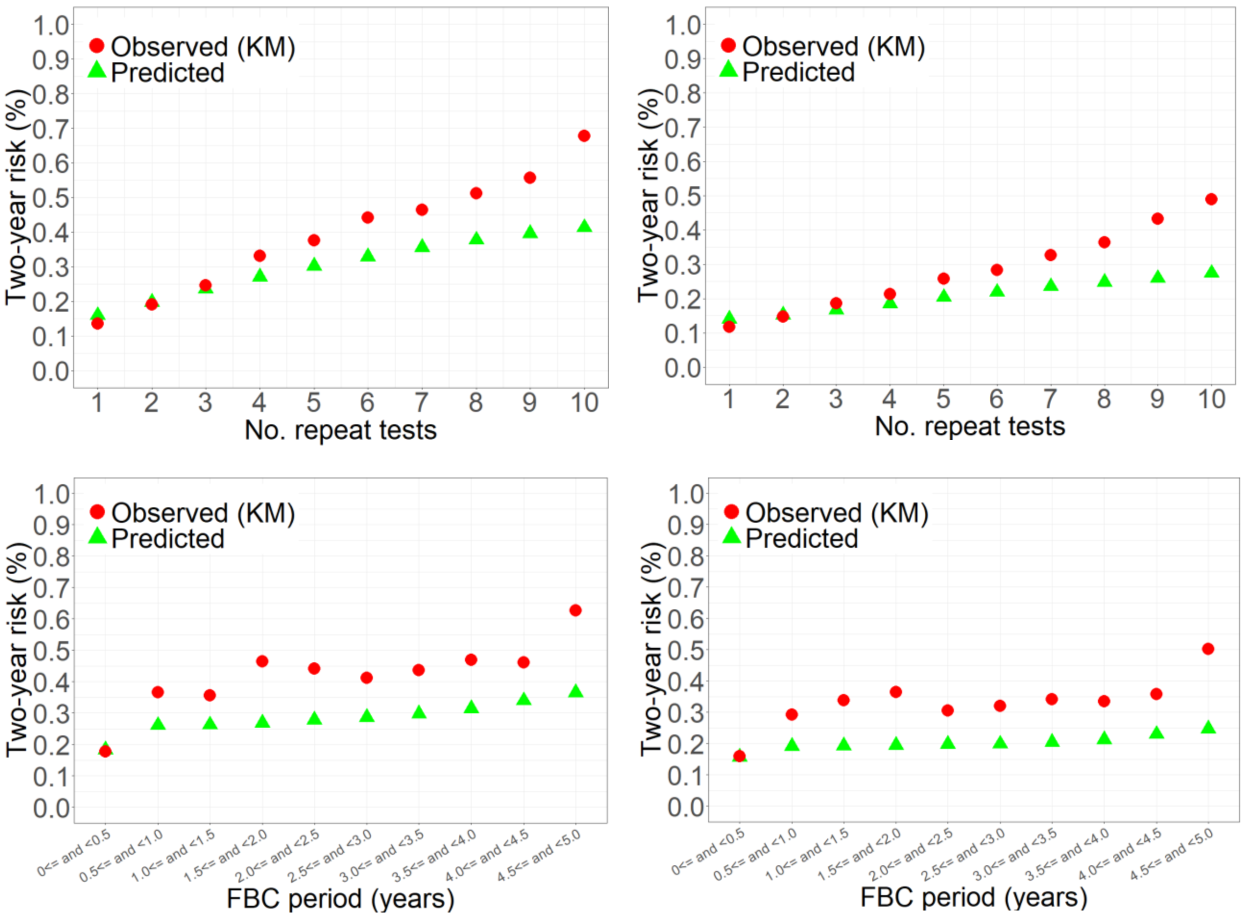

Abbreviations: KM=Kaplan-Meier

**Table S2: Diagnostic accuracy (95% CI) measures for males**

| Risk percentile | Risk threshold (%) | Sensitivity (%) | Specificity (%) | PPV (%) | NPV (%) |
|---|---|---|---|---|---|
| 1 | 0.0110 | 99.95 (99.95-99.95) | 1.00 (0.99-1.01) | 0.43 (0.42-0.44) | 99.98 (99.98-99.98) |
| 5 | 0.0152 | 99.73 (99.72-99.74) | 5.02 (4.99-5.04) | 0.45 (0.44-0.45) | 99.98 (99.98-99.98) |
| 10 | 0.0215 | 99.36 (99.35-99.37) | 10.04 (10.00-10.07) | 0.47 (0.46-0.48) | 99.97 (99.97-99.97) |
| 15 | 0.0298 | 98.79 (98.78-98.80) | 15.06 (15.02-15.10) | 0.49 (0.49-0.50) | 99.97 (99.96-99.97) |
| 20 | 0.0406 | 98.15 (98.13-98.16) | 20.07 (20.03-20.12) | 0.52 (0.51-0.53) | 99.96 (99.96-99.96) |
| 25 | 0.0542 | 97.19 (97.17-97.21) | 25.09 (25.05-25.14) | 0.55 (0.54-0.56) | 99.95 (99.95-99.95) |
| 30 | 0.0713 | 95.97 (95.95-95.99) | 30.11 (30.06-30.16) | 0.58 (0.57-0.59) | 99.94 (99.94-99.95) |
| 35 | 0.0929 | 94.29 (94.27-94.32) | 35.12 (35.07-35.18) | 0.62 (0.61-0.63) | 99.93 (99.93-99.93) |
| 40 | 0.1201 | 92.08 (92.05-92.11) | 40.14 (40.08-40.19) | 0.65 (0.64-0.66) | 99.92 (99.91-99.92) |
| 45 | 0.1533 | 89.10 (89.06-89.14) | 45.14 (45.09-45.20) | 0.69 (0.68-0.70) | 99.90 (99.89-99.90) |
| 50 | 0.1920 | 85.31 (85.27-85.35) | 50.15 (50.09-50.21) | 0.73 (0.72-0.74) | 99.88 (99.87-99.88) |
| 55 | 0.2344 | 80.59 (80.55-80.64) | 55.15 (55.09-55.21) | 0.76 (0.75-0.77) | 99.85 (99.85-99.85) |
| 60 | 0.2777 | 74.89 (74.84-74.94) | 60.15 (60.09-60.20) | 0.80 (0.79-0.81) | 99.82 (99.82-99.83) |
| 65 | 0.3195 | 68.22 (68.17-68.28) | 65.14 (65.09-65.20) | 0.83 (0.82-0.84) | 99.79 (99.79-99.80) |
| 70 | 0.3577 | 60.75 (60.69-60.80) | 70.13 (70.08-70.18) | 0.86 (0.85-0.87) | 99.76 (99.76-99.77) |
| 75 | 0.3926 | 52.60 (52.54-52.66) | 75.12 (75.07-75.17) | 0.89 (0.88-0.91) | 99.73 (99.73-99.74) |
| 80 | 0.4264 | 43.54 (43.49-43.60) | 80.10 (80.05-80.15) | 0.93 (0.92-0.94) | 99.70 (99.69-99.71) |
| 85 | 0.4626 | 34.23 (34.17-34.28) | 85.08 (85.04-85.12) | 0.97 (0.96-0.98) | 99.67 (99.66-99.68) |
| 90 | 0.5079 | 24.30 (24.26-24.35) | 90.06 (90.03-90.09) | 1.03 (1.02-1.05) | 99.64 (99.64-99.65) |
| 95 | 0.5839 | 13.63 (13.59-13.67) | 95.04 (95.01-95.06) | 1.16 (1.15-1.17) | 99.61 (99.61-99.62) |
| 99 | 0.7810 | 3.56 (3.54-3.58) | 99.01 (99.00-99.02) | 1.52 (1.50-1.53) | 99.59 (99.58-99.59) |

**Table S3: Diagnostic accuracy (95% CI) measures for females**

| Risk percentile | Risk threshold (%) | Sensitivity (%) | Specificity (%) | PPV (%) | NPV (%) |
|---|---|---|---|---|---|
| 1 | 0.0084 | 99.88 (99.88-99.89) | 1.00 (0.99-1.01) | 0.34 (0.33-0.34) | 99.96 (99.96-99.96) |
| 5 | 0.0120 | 99.48 (99.47-99.49) | 5.01 (4.99-5.03) | 0.35 (0.34-0.36) | 99.97 (99.96-99.97) |
| 10 | 0.0168 | 98.88 (98.87-98.89) | 10.02 (9.99-10.06) | 0.37 (0.36-0.37) | 99.96 (99.96-99.96) |
| 15 | 0.0228 | 98.18 (98.17-98.20) | 15.04 (15.01-15.08) | 0.39 (0.38-0.39) | 99.96 (99.96-99.96) |
| 20 | 0.0304 | 97.30 (97.29-97.32) | 20.06 (20.02-20.10) | 0.41 (0.40-0.41) | 99.95 (99.95-99.96) |
| 25 | 0.0400 | 96.35 (96.33-96.37) | 25.07 (25.03-25.12) | 0.43 (0.42-0.44) | 99.95 (99.95-99.95) |
| 30 | 0.0521 | 94.94 (94.92-94.96) | 30.08 (30.04-30.13) | 0.45 (0.45-0.46) | 99.94 (99.94-99.95) |
| 35 | 0.0676 | 93.58 (93.55-93.60) | 35.10 (35.05-35.14) | 0.48 (0.48-0.49) | 99.94 (99.94-99.94) |
| 40 | 0.0878 | 91.47 (91.44-91.50) | 40.11 (40.05-40.16) | 0.51 (0.50-0.52) | 99.93 (99.93-99.93) |
| 45 | 0.1133 | 88.95 (88.92-88.98) | 45.11 (45.06-45.17) | 0.54 (0.53-0.55) | 99.92 (99.91-99.92) |
| 50 | 0.1434 | 85.96 (85.93-86.00) | 50.12 (50.07-50.17) | 0.58 (0.57-0.58) | 99.91 (99.90-99.91) |
| 55 | 0.1750 | 82.08 (82.04-82.12) | 55.12 (55.07-55.18) | 0.61 (0.60-0.62) | 99.89 (99.89-99.89) |
| 60 | 0.2057 | 76.94 (76.90-76.98) | 60.12 (60.07-60.17) | 0.64 (0.64-0.65) | 99.87 (99.87-99.87) |
| 65 | 0.2342 | 70.99 (70.94-71.03) | 65.12 (65.07-65.17) | 0.68 (0.67-0.69) | 99.85 (99.85-99.85) |
| 70 | 0.2605 | 63.93 (63.88-63.98) | 70.11 (70.07-70.16) | 0.71 (0.71-0.72) | 99.83 (99.82-99.83) |
| 75 | 0.2857 | 56.65 (56.59-56.70) | 75.11 (75.06-75.15) | 0.76 (0.75-0.77) | 99.81 (99.80-99.81) |
| 80 | 0.3122 | 48.29 (48.24-48.34) | 80.09 (80.05-80.14) | 0.81 (0.80-0.82) | 99.78 (99.78-99.79) |
| 85 | 0.3429 | 38.87 (38.82-38.92) | 85.08 (85.04-85.12) | 0.87 (0.86-0.88) | 99.76 (99.75-99.76) |
| 90 | 0.3840 | 28.93 (28.88-28.98) | 90.06 (90.03-90.09) | 0.97 (0.96-0.98) | 99.74 (99.73-99.74) |
| 95 | 0.4579 | 17.06 (17.02-17.10) | 95.04 (95.02-95.06) | 1.14 (1.13-1.15) | 99.71 (99.70-99.71) |
| 99 | 0.6872 | 5.03 (5.00-5.05) | 99.01 (99.00-99.02) | 1.68 (1.67-1.70) | 99.68 (99.67-99.68) |

**Table S4: Summary of cancer diagnosis by age group**

| Cancer (n (%)) | Main analysis (NCRAS cancers) | | Sensivity analysis (NCRAS+CPRD+HES+ONS cancers) | |
|---|---|---|---|---|
| | **Males** | **Females** | **Males** | **Females** |
| Overall | 12,578 (0.4%) | 11,939 (0.3%) | 13,423 (0.5%) | 12,919 (0.4%) |
| Age (years) at baseline test | | | | |
| Age 18-39 | | | | |
| Age 40-49 | 328 (0.1%) | 446 (0.1%) | 375 (0.1%) | 495 (0.1%) |
| Age 50-59 | 1,265 (0.2%) | 1,018 (0.1%) | 1,352 (0.2%) | 1,125 (0.1%) |
| Age 60-69 | 3,357 (0.5%) | 2,191 (0.3%) | 3,553 (0.6%) | 2,341 (0.3%) |
| Age 70-79 | 4,628 (0.9%) | 4,006 (0.7%) | 4,920 (0.9%) | 4,273 (0.7%) |
| Age 80-89 | 2,723 (0.9%) | 3,704 (0.8%) | 2,919 (0.9%) | 4,018 (0.8%) |
| Age 90+ | 277 (0.6%) | 574 (0.4%) | 304 (0.6%) | 667 (0.5%) |

**Figure S10: Calibration plots for the BLOODTRACC models – sensitivity analysis (NCRAS+CPRD+HES+ONS cancers)**

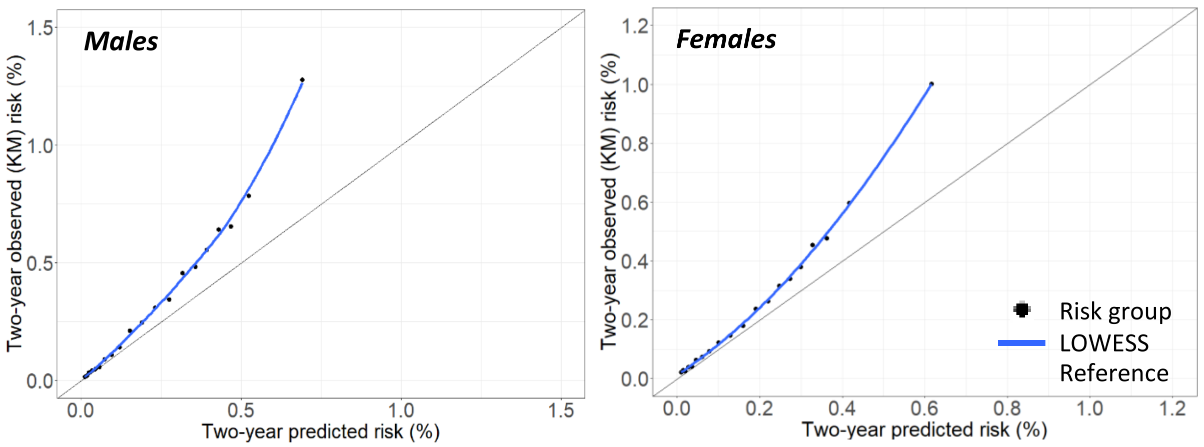

**Table S5: Summary of cancer diagnosis by study design and age group**

| Cancer (n (%)) | Males | | Females | |
|---|---|---|---|---|
| | **Main analysis (hybrid)** | **Sensitivity analysis (cohort)** | **Main analysis (hybrid)** | **Sensitivity analysis (cohort)** |
| Overall | 12,578 (0.4%) | 18,876 (0.4%) | 11,939 (0.3%) | 17,501 (0.3%) |
| Age (years) at baseline test | | | | |
| Age 40-49 | 328 (0.1%) | 795 (0.1%) | 375 (0.1%) | 724 (0.1%) |
| Age 50-59 | 1,265 (0.2%) | 2,211 (0.3%) | 1,352 (0.2%) | 1,617 (0.2%) |
| Age 60-69 | 3,357 (0.5%) | 4,217 (0.6%) | 3,553 (0.6%) | 2,614 (0.4%) |
| Age 70-79 | 4,628 (0.9%) | 5,703 (0.9%) | 4,920 (0.9%) | 4,470 (0.7%) |
| Age 80-89 | 2,723 (0.9%) | 4,668 (1.1%) | 2,919 (0.9%) | 5,766 (0.9%) |
| Age 90+ | 277 (0.6%) | 893 (0.9%) | 304 (0.6%) | 1,886 (0.8%) |
